# SIGLEC1 (CD169): a marker of active neuroinflammation in the brain but not in the blood of MS patients

**DOI:** 10.1101/2020.12.18.20248500

**Authors:** Lennard Ostendorf, Philipp Dittert, Robert Biesen, Ankelien Duchow, Victoria Stiglbauer, Klemens Ruprecht, Dominik Seelow, Raluca A. Niesner, Anja E. Hauser, Friedemann Paul, Helena Radbruch

**Author notes:** Corresponding Authors: Lennard Ostendorf, Helena Radbruch.

## Abstract

**Objective:** We aimed to evaluate SIGLEC1 (CD169) as a biomarker in Multiple Sclerosis (MS) and Neuromyelitis optica spectrum disorder (NMOSD) and to evaluate the specificity of SIGLEC1^+^ myeloid cells for demyelinating diseases

**Methods:** We performed flow cytometry-based measurements of SIGLEC1 expression on monocytes in 86 MS patients, 41 NMOSD patients and 31 healthy controls. Additionally, we histologically evaluated the presence of SIGLEC1+ myeloid cells in acute and chronic MS brain lesions as well as other neurological diseases.

**Results:** We found elevated SIGLEC1 expression in 16/86 (18.6%) MS patients and 4/41 (9.8%) NMOSD patients. Almost all MS patients with high SIGLEC1 levels received exogenous interferon beta as an immunomodulatory treatment and only a small fraction of MS patients without interferon treatment had increased SIGLEC1 expression. SIGLEC1+ myeloid cells were abundantly present in active MS lesions as well as in a range of acute infectious and malignant diseases of the central nervous system, but not chronic MS lesions.

**Conclusion:** In our cohort, SIGLEC1 expression on monocytes was – apart from those patients receiving interferon treatment – not significantly increased in patients with MS and NMOSD, nor were levels associated with more severe disease. The presence of SIGLEC1+ myeloid cells in brain lesions could be used to investigate the activity in an inflammatory CNS lesion.

## Background

Multiple sclerosis (MS) and neuromyelitis optica spectrum disorder (NMOSD) are both chronic, demyelinating inflammatory diseases of the central nervous system that are a significant cause of disability in the young. While certain aspects of the pathophysiology of both diseases have been uncovered, many aspects towards a complete understanding remain to be elucidated.^1^

SIGLEC1 (Sialic acid-binding immunoglobulin-type lectins-1, CD169) is a sialic acid binding cell-surface protein, exclusively expressed on monocytes and macrophages.^2^ Its expression is upregulated upon contact with type I interferons and to a lesser degree, other activatory stimuli such as LPS.^3,4^ As such, SIGLEC1 expression has been used as a surrogate marker for type I interferon activity in autoimmune diseases like Systemic lupus erythematosus^5^ or primary Sjögren syndrome^6^ as well as interferonopathies^7^ and viral infections^8,9^.

In the healthy brain, SIGLEC1 is only expressed by some perivascular and choroid plexus macrophages, but not by microglia.^10^ In mice, mechanical insult to the brain led to an accumulation of SIGLEC1+ myeloid cells in the damaged area.^10^ The cause of this accumulation, however is unclear – either the contact of serum components or inflammatory signals with resident myeloid cells induces SIGLEC1 expression or blood-derived SIGLEC1+ myeloid cells infiltrate the CNS.

The role of type I interferons and SIGLEC1 in the pathophysiology of MS and NMOSD is not yet clear. In relapsing-remitting MS (RRMS), type I interferons are used as an immunomodulatory treatment that reduces the rate of relapses.^11^ It does, however, not protect against clinical progression in the progressive forms of MS (primary progressive MS (PPMS) and secondary progressive MS (SPMS)).^11^ In a recent report, SIGLEC1 positive myeloid cells were found within MS lesions and specific ablation of SIGLEC1 expressing cells in the experimental autoimmune encephalitis (EAE) model of MS led to an amelioration of disease.^12^ Additionally, plasmacytoid dendritic cells, the main type I interferon-producing cells, were found in increased frequencies in the cerebrospinal fluid of MS patients during a flare.^13^ Two studies found SIGLEC1 expression to be increased on blood monocytes of MS patients, especially those with a progressive type of MS.^12,14^ Taken together, there is conflicting evidence on the role of type I interferons and the interferon-induced expression of SIGLEC1 in MS as being protective or pathogenic. This is likely due to the heterogeneity of patients and disease stages and potentially also the confounding effect of interferon therapy.

In NMOSD, little is known about the role of type I interferons; however, many patients with NMOSD have an overlap with additional, type I interferon-dependent diseases, such as SLE^15^ and a recent report describes a series of patients with increased levels of endogenous or exogenous interferon α who went on to develop a seropositive NMOSD.^16^ Interferon treatment does not prevent NMOSD relapses^17^ and anecdotally even increases the disease activity.^18^ Additionally, we recently described the presence of low-density granulocytes in MS and NMOSD patients,^19^ a subset of granulocytes that produce high levels of type I interferons in SLE.^20^ Thus, we hypothesized that type I interferons could also play a role in the pathogenesis of NMOSD.

In this work we investigated expression of SIGLEC1 on monocytes of patients with MS and NMOSD and correlated the expression with clinical parameters. In addition, we analysed human brain tissue sections of patients with both active and inactive MS as well as other neurological and systemic diseases.

## Methods

### Cohort

We analysed frozen peripheral blood mononuclear cells (PBMCs) from a total of 86 MS patients, 41 NMOSD patients and 31 healthy controls (HCs) that were included in observational cohort studies for MS and NMOSD at the NeuroCure Clinical ResearchCenter, Charité—Universitätsmedizin Berlin. These studies were approved by the ethics committee of the Charité (MS: EA1/163/12; NMOSD: EA1/041/14) and informed consent was obtained from all participants in the study. PBMCs from all patients with sufficient available material were included in the study with no prior selection according to disease activity, treatment, age or sex. All MS patients fulfilled the 2017 revised McDonald criteria,^21^ while the NMOSD patients fulfilled the 2015 Wingerchuck international consensus diagnostic criteria.^22^ Further patient characteristics are provided in table 1.

**Table 1:** Epidemiological Data. (IQR: Interquartile range; RRMS: Relapse-Remitting Multiple Sclerosis; SPMS: Secondary Progressive Multiple Sclerosis; PPMS: Primary Progressive Multiple Sclerosis; AQP4+: Aquaporin-4 antibody positive; MOG+: Myelin oligodendrocyte glycoprotein antibody positive; MCTD: Mixed connective tissue disease)

|  | Healthy Controls | MS Patients | NMOSD Patients |
| --- | --- | --- | --- |
| Number of Individuals | 31 | 86 | 41 |
| Age (median, IQR) | 31 (27 – 34) | 45 (34 – 52) | 57 (45 – 64) |
| Sex (n female) | 15 (48.4%) | 53 (61.6%) | 37 (90.2%) |
| Subtype (n) | – | RRMS: 63 (73.3%)<br>SPMS: 15 (17.4%)<br>PPMS: 5 (5.9%) | AQP4+: 23 (56.1%)<br>MOG+: 7 (17.1%)<br>seronegative: 3 (7.3%)<br>unknown: 8 (19.5%) |
| Additional Immune-mediated Diseases (n) | Autoimmune Thyroiditis (2),<br>Asthma bronchiale (1) | Autoimmune Thyroiditis (10), Asthma bronchiale (2), Psoriasis (1), Idiopathic Myocarditis (1), Celiac Disease (1) | Autoimmune Thyroiditis (3), Myasthenia gravis (1), Sjögren Syndrome (1), MCTD (1) |
| Immunosuppressive Medication (n) | – | None: 24 (27.9%)<br>Dimethyl fumarate: 15 (17.4%),<br>Glatiramer acetate: 14 (16.3%),<br>Interferon beta: 13 (15.1%),<br>Fingolimod: 7 (8.1%),<br>Teriflunomide: 5 (5.8%),<br>Methylprednisolone: 2 (2.3%),<br>Daclizumab: 2 (2.3%),<br>Rituximab: 1 (1.2%),<br>Ocrelizumab: 1 (1.2%),<br>Natalizumab 1 (1.2%) | Rituximab: 19 (46.3%)<br>Azathioprine: 9 (22.0%)<br>None: 6 (14.6%)<br>Prednisolone: 3 (7.3%)<br>Mycophenolate: 2 (4.9%)<br>Glatiramer acetate: 1 (2.4%)<br>unknown: 1 (2.4%) |

Longitudinal samples were available for 28/86 MS patients, 12/41 NMOSD patients and 12/31 healthy controls. Follow-up periods were up to 1965 days and included 2 to 5 time points.

### PBMC isolation and flow cytometry

Peripheral blood mononuclear cells (PBMCs) were isolated from EDTA anticoagulated blood. Blood and phosphate-buffered saline supplemented with 0.5% bovine serum albumin (PBS/BSA) were mixed at a 1:1 ratio and 35 ml were layered onto 15 ml Ficoll-Paque PLUS gradient (GE Healthcare). PBMCs were isolated according to the manufacturer’s protocol and stored in liquid nitrogen until the analysis. For analysis batches of 20-35 samples were thawed and washed in RPMI 1640 cell medium (Gibco). In each batch, a mix of samples from healthy controls and MS/NMOSD patients were included. Cells were stained with antibodies against CD14 (DRFZ, clone TM1), CD169 (SIGLEC1, clone 7-239, BioLegend Cat# 346004, RRID:AB_2189029) and a dead cell stain (eBioscience Fixable Viability Dye eFluor™ 780). For each batch, a fluorescence minus one (FMO) control for SIGLEC1 was measured and FMO measurements remained stable over multiple batches. The samples were acquired on a FACSCanto cytometer (BD Biosciences) and all cytometry experiments were performed according to published standards.^23^ Using the FlowJo software (Version 10.4.1 for Mac, FlowJo LLC), we identified monocytes and excluded doublets based on scatter parameters. We then gated on CD14^high^, living cells and analysed the median fluorescence intensity (MFI) for SIGLEC1. (Fig. 1a)

**Figure 1:**
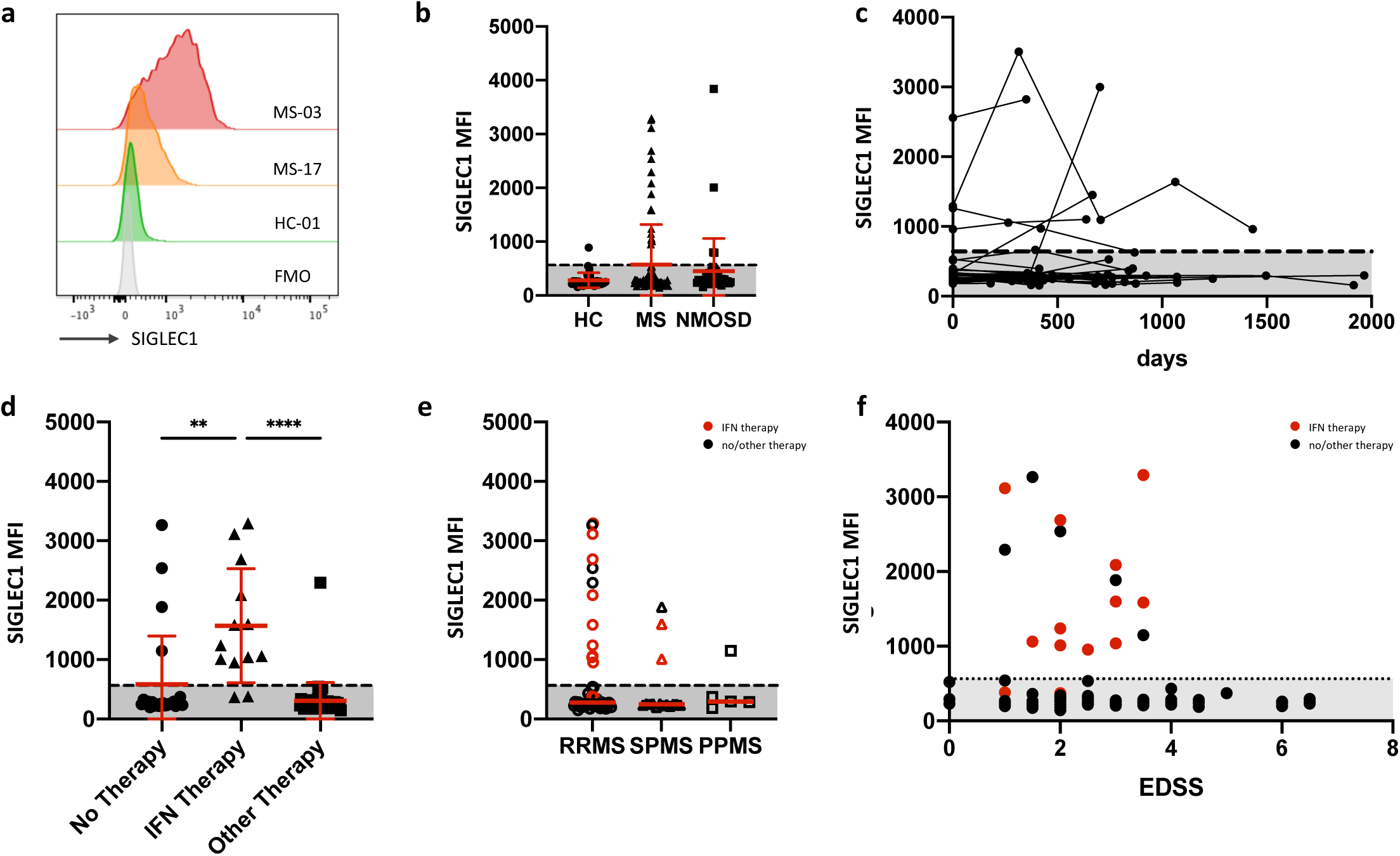
(a) Representative histograms of SIGLEC1 fluorescence on CD14^high^ monocytes, FMO: fluorescence minus one control. (b) Cross-sectional analysis of SIGLEC1 expression on CD14^high^ monocytes of 31 healthy controls, 86 MS patients and 41 NMOSD patients. If multiple samples of the same individual were analysed, the mean of the measurements is displayed. (c) Longitudinal analysis of SIGLEC1 expression of 52 individuals with up to five measurements. (d) Comparison of SIGLEC1 expression in MS patients, receiving no immunomodulatory treatment (n = 24), interferon beta (n = 13) or a non-interferon treatment (n = 49). Treatment with interferon with significantly associated with higher levels of SIGLEC1 expression: Kruskal-Wallis test with Dunn’s correction, **: p<0.01, ****: p<0.0001. (e) Comparison of SIGLEC1 expression in patients with RRMS (n = 63), SPMS (n = 15) and PPMS (n = 5). Red symbols indicate interferon treatment. (f) Scatter plot of the EDSS disability score against SIGLEC1 expression, no significant correlation was detected.

### Histology of human brain tissue

We investigated archived cryo- and formalin preserved biopsy and autopsy tissue from patients who had been diagnosed in the Department of Neuropathology, Charité – Universitätsmedizin Berlin with inflammatory demyelination of the central nervous system (CNS) consistent with multiple sclerosis or other inflammatory, malignant or infectious diseases of the CNS (other neurological disease, OND). Patients who died of cardiovascular cases or multi-organ failure served as controls. All patients with multiple sclerosis fulfilled clinical diagnosis criteria according to the 2017 revised McDonald criteria13 and none of the patients were treated with interferon.

### Immunohistology of human brain tissue

All stains were performed on 8μm cryomicrotome sections, cut on a NX80 cryotome (ThermoFisher, Waltham, Massachusetts, USA) according to standard procedures. Stains included Hematoxylin and eosin (H&E), CD68 (EBM11, Dako #M0718, 1:100), HLA-DR (CR3/43; Dako #M0775, 1:200) and SIGLEC1 (HSn7D2, Novus Biologicals, 1:200). Immunohistochemical stainings were performed on a Benchmark XT autostainer (Ventana Medical Systems, Tuscon, AZ, USA). The presence of SIGLEC1 perivascular and leptomeningeal staining served as a quality control. Microscopy was performed on a BZ-9000 BioRevo microscope (Keyence, Neu-Isenburg, Germany). Positively stained cells were defined by presence of a nucleus and cytoplasmic staining.

### Data analysis

Statistical analyses were performed using Python 3.7.6 with the Numpy (v1.19.0)^24^ and Pandas (v1.1.0)^25^ packages as well as GraphPad Prism (v8.4.3. for Mac). A normal range for SIGLEC1 expression in FACS expression was calculated as two standard deviations from the mean SIGLEC1 expression in healthy controls. When multiple measurements of the same individual were available, the mean of the measurements was calculated and used for comparative analysis. The Kruskal-Wallis test with Dunn’s correction was used to compare between the different groups.

## Results

### SIGLEC1 expression is increased in a subset of patients with multiple sclerosis

We investigated the SIGLEC1 expression on CD14^+^ monocytes in the peripheral blood of MS and NMOSD patients as well as in healthy controls. In most of the samples, monocyte SIGLEC1 expression was barely above the negative (fluorescence minus one) control (Fig. 1a, b). We defined a physiologic range of SIGLEC1 expression as two standard deviations from the mean SIGLEC1 expression in healthy controls (normal MFI range 0 – 564). Accordingly, 16/86 (18.6%) MS patients, 4/41 (9.8%) NMOSD patients and 1/31 (3.2%) healthy controls had increased SIGLEC1 levels. Almost all individuals with available longitudinal samples remained in their respective SIGLEC1 high or low category over a follow-up period of up to 1965 days. (Fig. 1c)

### Correlation with clinical parameters

Next, we aimed to identify parameters that were associated with increased SIGLEC1 in MS patients. One obvious explanation would be exogenous type 1 interferon as treatment. 11/16 MS patients (68.6%) with increased SIGLEC1 expression levels received interferon beta treatment at the time of measurement (Fig. 1d). While a previous report found increased SIGLEC1 levels to be more prevalent in MS patients with a progressive form of MS, we found only 2/20 patients primary or secondary progressive MS with increased SIGLEC1 levels that were not explained by interferon treatment (Fig. 1e). SIGLEC1 expression on monocytes did not correlate with the Expanded Disability Status Scale (EDSS) (Fig. 1f), nor had it a temporal association with relapses.

### SIGLEC1 expression on brain-infiltrating myeloid cells

To investigate the presence and specificity of SIGLEC1+ myeloid cells in inflammatory MS lesion, we analysed brain tissue sections from 4 patients with active relapsing-remitting MS (RRMS), 5 patients with secondary-progressive MS (SPMS) and 6 patients who died of cardiovascular or multi-organ failure. In all control samples, SIGLEC1 positivity was limited to cells in the leptomeninges as well as perivascular macrophages, as previously reported.^10^ In all four samples from patients with active relapsing-remitting MS (RRMS) as well as in one patient with the Marburg variant of MS, we found a dense infiltrate of CD68+HLA-DR+ myeloid cells that stained predominantly positive for SIGLEC1 (Fig. 2a). In five samples from patients with secondary-progressive MS (SPMS) however, we found CD68+HLA-DR+ infiltrates of varying density, but almost no SIGLEC1 expression (Fig. 2a). This indicates that SIGLEC1 expression on myeloid cells could be used to distinguish active inflammatory lesions from chronic lesions.

**Figure 2:**
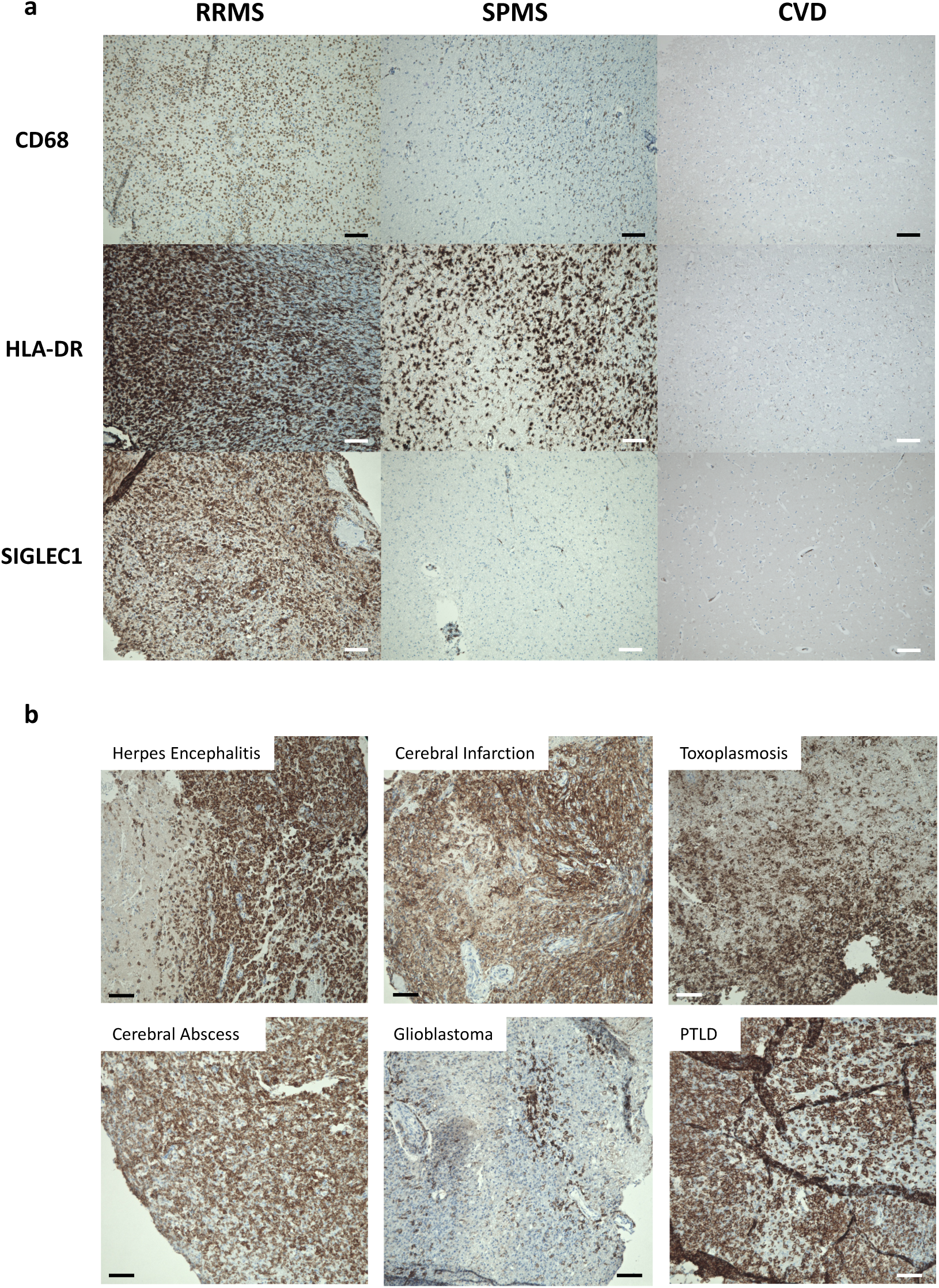
(a) Representative immunohistochemical stainings of CD68, HLA-DR and SIGLEC1 (CD169) in brain tissue from a patient with active relapsing-remitting multiple sclerosis (RRMS), secondary-progressive multiple sclerosis (SPMS) and a control patient who died of cardiovascular disease (CVD). Both RRMS and SPMS show infiltrates with CD68+ and HLA-DR+ myeloid cells, but only in RRMS, these express SIGLEC1. Nuclei are stained blue and the respective marker antigens in brown. The scale bar indicates 100µm. These results are representative of 4 RRMS, 5 SPMS and 7 control samples that were examined. (b) Immunohistocheminal stainings of SIGLEC1 in brain tissue from patients with Herpes simplex viral encephalitis (n=2), cerebral infarction with inflammatory changes (n=2), cerebral toxoplasmosis (n=1), cerebral abscess (n=2), glioblastoma (n=2) and post-transplant lymphoproliferative disease (PTLD, n=1). Infiltrates of SIGLEC1+ myeloid cells were observed in all samples.

To corroborate this hypothesis, we studied the SIGLEC1 expression in the brain tissue of 8 patients with other inflammatory neurological diseases. SIGLEC1+ myeloid infiltrates were present in patients with glioblastoma (n=2), herpes simplex encephalitis (n=2) and cerebral infarction (n=2), as well as toxoplasmosis (n=1), cerebral abscess (n=2) and post-transplant lymphoproliferative disorders (n =1, PTLD, Fig. 2b). No increase in SIGLEC1+ cells was noted in the brain of a patient who died of amyotrophic lateral sclerosis (ALS).

In summary, SIGLEC1+ is expressed on brain-infiltrating myeloid cells in a broad range of active inflammatory lesions, but not in chronic MS lesions.

## Conclusion

Here, we report that, after accounting for interferon treatment, patients with multiple sclerosis (MS) and neuromyelitis optica spectrum disorder (NMOSD) did not have increased expression of SIGLEC1 on monocytes in the peripheral blood.

We identified SIGLEC1+ on CD68+HLA-DR+ myeloid cells in active inflammatory MS lesions and a range of other inflammatory, infectious or malignant brain lesions. SIGLEC1+ expression was low on myeloid cells in chronic MS lesions of SPMS patients, indicating that SIGLEC1+ myeloid cells could serve as a marker of inflammatory activity. These findings are in line with a previous report that found SIGLEC1+MHC-II+ myeloid cells to be present in mice after retinal transplantation, but not in the healthy retina or retinal degeneration.^26^ Future studies will be needed to investigate, whether the increase in SIGLEC1+ cells in inflammatory brain lesions is due to the infiltration of SIGLEC1+ cells from the peripheral blood or an upregulation of SIGLEC1 in tissue-resident microglia, as well as define the inflammatory signals leading to such an upregulation. We propose that SIGLEC1+ can serve as a marker to differentiate active from inactive MS lesions, which will require additional validation for use in routine histopathological diagnostics.

Our results on SIGLEC1 expression on blood monocytes are in contrast to two previous reports which identified increased levels of SIGLEC1 in smaller cohorts of MS patients.^12,14^ The study by Malhotra et al^14^ focussed on untreated MS patients and identified modest increases in SIGLEC1 expression in MS patients and especially those with progressive disease. A second study by Bogie et al^12^ again identified small increases in SIGLEC1 expression on monocytes in MS patients and found that these increases were independent of progressive disease and also of interferon treatment, an observation that is at odds with our own findings and biological plausibility.

This disparity might be due to different reasons: it could be that the authors mostly observed small differences between individuals that would have all been classified as “SIGLEC1 low expressors” in our study, as the fold-differences they describe are significantly smaller compared to the 5-10-fold increases in “SIGLEC1 high expressors” in our study. Different studies investigated the SIGLEC1 expression using different antibody clones which could also explain some of the differences; in this study, we used the same clone as previous studies that investigated SIGLEC1 in rheumatologic diseases.^5,6^ One significant weakness of our own study is the relative clinical quiescence of the patients enrolled; most did not have a relapse within the last six months before the measurement and most did not have a change in EDSS in longitudinal sampling. Additionally, most patients were under immunosuppressive treatment. Future studies in MS and NMOSD patients during a clinical relapse could thus still discover the presence of SIGLEC1 expressing monocytes in the peripheral blood. In summary, our data indicate that type I interferons or SIGLEC1 expressing cells in the peripheral blood do not play a major role in the pathogenesis of most patients with stable NMOSD or MS, however SIGLEC1^+^ myeloid cells in the brain are present in inflammatory MS lesions as well as in other inflammatory neurological diseases of the CNS.

## Data Availability

All datasets used and analysed during the current study are available from the corresponding authors on request.

## Declarations

### Ethics approval and consent to participate

These studies were approved by the ethics committee of the Charité – Universitätsmedizin Berlin (references: EA1/163/12, EA1/041/14, EA1/107/13) and informed consent was obtained from all participants in the study.

### Consent for publication

Not applicable.

